# Use of the Wnt/β-catenin Activator Lithium Is Associated with Less Emphysema

**DOI:** 10.1101/2025.07.24.25332150

**Authors:** Divay Chandra, Kassandra Allbright, Gregory L Kinney, Yisha Li, Russell Bowler, Karina Serban, Toru Nyunoya, Puja Dutta, Stephen Rennard, Frank Sciurba, Melanie Königshoff

## Abstract

**Rationale:** Emphysema is defined by progressive alveolar destruction and impaired tissue repair, with diminished Wnt/β-catenin signaling implicated in its pathogenesis. Preclinical studies suggest that lithium, a pharmacologic activator of Wnt/β-catenin signaling, may attenuate emphysema. However, its effects in humans remain unknown.

**Objectives:** To investigate whether individuals using lithium for neuropsychiatric conditions have reduced susceptibility to emphysema compared to users of other neuropsychiatric medications.

**Methods:** We analyzed cross-sectional data from two large cohorts - the UK Biobank and COPDGene - comprising over 800 individuals using oral lithium. Lithium users were compared to individuals using other neuropsychiatric medications. In the UK Biobank, outcomes included spirometry and self-reported physician-diagnosed emphysema. In COPDGene, outcomes included spirometry and quantitative CT measures of emphysema. Multivariable regression and propensity score matching accounted for demographics, smoking history, and psychiatric diagnoses.

**Measurements and Main Results:** In the UK Biobank, lithium use was associated with higher FEV₁ and FVC (% predicted) and ∼50% lower adjusted odds of emphysema diagnosis. In COPDGene, lithium users exhibited significantly higher FEV₁, FVC, and FEV₁/FVC ratios, lower CT-measured emphysema (%LAA-950), and higher lung density. These associations persisted after multivariable adjustment and across sensitivity analyses.

**Conclusions:** Lithium use is associated with less emphysema in two independent cohorts. These findings align with preclinical evidence supporting Wnt/β-catenin activation as a protective mechanism and warrant further investigation of lithium and related agents as potential therapies for emphysema.

## INTRODUCTION

Chronic obstructive pulmonary disease (COPD) is a leading cause of death and disability worldwide. Despite the substantial clinical burden, there are currently no disease-modifying therapies for COPD. Emphysema is a key pathological feature of COPD, contributing substantially to its morbidity and mortality. Emphysema is characterized by progressive damage and destruction of alveolar structures. Wnt/β-catenin signaling is essential for alveolar repair; however, prior studies, including our own, have shown reduced Wnt/β-catenin signaling in emphysematous lungs compared to controls (1–6). These findings suggest that impaired Wnt/β-catenin signaling contributes to emphysema pathogenesis and may represent a promising therapeutic target for this disease.

Lithium, a mood stabilizing medication, is a well-established activator of the Wnt/β-catenin pathway. Wnt/β-catenin signaling is initiated when extracellular Wnt ligands bind to the Frizzled family of cell surface receptors (7). Ligation of these receptors prevents a multi-protein complex called the β-catenin destruction complex from degrading β-catenin in the cytosol. As a result, β-catenin accumulates in the cytosol and moves to the nucleus, where it triggers transcription of Wnt target genes that regulate cell proliferation, survival, and differentiation. Lithium activates Wnt/β-catenin signaling by inhibiting GSK3β, a crucial component of the β-catenin destruction complex (8, 9).

We previously demonstrated that lithium upregulates transcripts and proteins involved in Wnt/β-Catenin signaling, increases surfactant protein C (SFTPC) expression in alveolar type 2 cells, and promotes linear elastin deposition along alveolar walls in COPD patient-derived precision-cut lung slices compared to untreated tissue (2).

Enhanced SFTPC expression and elastin deposition are key reparative processes in the lung (10–13). We further demonstrated that lithium administration attenuates emphysema, increases epithelial cell marker expression, suppresses matrix metalloproteinase-12 (MMP-12), and modulates macrophage function and elastin remodeling in both murine models and human 3D *ex vivo* lung tissue (1, 2).

Based on the accumulating evidence, we hypothesized that individuals receiving lithium for neuropsychiatric conditions would exhibit reduced emphysema. To test this hypothesis, we analyzed data from two large independent, well-characterized cohorts: the UK Biobank and COPDGene.

## METHODS

### UK Biobank Cohort

The UK Biobank cohort includes approximately 500,000 participants (Application Number 60327), as previously described (14). Established to investigate the genetic and environmental determinants of various diseases, the cohort enrolled individuals aged 40– 69 years from diverse geographic, socioeconomic, and ethnic backgrounds across the United Kingdom between 2006 and 2010. Participants underwent comprehensive assessments, including questionnaires regarding comorbidities, pre-bronchodilator spirometry, and physical and functional measurements.

Medication use was self-reported. Data on the use of lithium and other mood-stabilizing medications at the baseline visit were extracted from the UK Biobank medication inventory, as previously detailed (15). In brief, lithium users were compared with those using Olanzapine, Quetiapine, Risperidone, Aripiprazole, Haloperidol, Amisulpride, Chlorpromazine, Flufentixol, Sulpiride, Biquelle, Trifluoperazine, Promazine, Stelazine, Mintreleq, Depakote, Valproic Acid, Pericyazine, Zuclopenthixol, or Zaluron (15). Pre-bronchodilator spirometry was assessed in the cohort and used to calculate percent predicted values as described previously (16).

Baseline demographics, clinical characteristics, and lung function measures were compared between lithium users and users of other mood-stabilizing medications using descriptive statistics. Multivariable logistic regression models were constructed to assess the association between lithium use (independent variable: yes/no) and physician-diagnosed emphysema (dependent variable: yes/no), adjusting for age, sex, race/ethnicity, body mass index (BMI), and self-reported diagnoses of mania, bipolar disorder, schizophrenia, or schizoaffective disorder. Among ever smokers, models were further adjusted for cumulative tobacco exposure (pack-years of smoking).

### COPDGene Cohort

Enrollment criteria for the COPDGene cohort have been described previously (17, 18). In brief, participants were 45-80 years old with >10 pack-years of current or prior smoking, without prior thoracic surgery or lung disease besides asthma or COPD. Because the presence of airflow obstruction, i.e., FEV_1_/FVC<0.70, was not required for enrollment, the study included smokers with and without COPD. The Institutional Review Boards at all participating sites approved the study. Written informed consent was obtained from each participant.

Demographics, educational attainment, smoking history, and medication use were self-reported at enrollment. Participants who reported oral lithium use for any indication were classified as lithium users.

CT images were acquired at full inspiration and analyzed to assess emphysema by density mask approach (low-attenuation area using a −950 HU threshold or LAA %F950) and to measure adjusted lung density (grams/Liter), as described previously (19). Post-bronchodilator spirometry was performed and adjusted to standard population-derived predicted values (20).

Baseline characteristics, CT-based measures of emphysema, and spirometry were compared between lithium users and users of other neuropsychiatric medications. Multivariable linear regression models were constructed to assess the association between lithium use (independent variable: yes/no) and radiographic measures of emphysema - LAA %F950 and average lung density (dependent variables) – while adjusting for age, gender, race/ethnicity, educational attainment, and pack-years of smoking as covariates. LAA% −950 was log-transformed because of its right-skewed distribution.

To further address potential confounding, lithium users were propensity-matched to non-users in a 1:2 ratio using the “teffects” package in Stata MP version 18, with a caliper of 0.03 (StataCorp, College Station, TX). Covariates for the propensity model were chosen according to the recommendations of Brookhart et al. (19). Multiple sensitivity analyses were performed to assess the robustness of the findings, including nearest neighbor matching with Mahalanobis distance, as well as alternative causal inference approaches such as regression adjustment, inverse probability weighting (IPW), and double robust methods (IPW with regression adjustment).

All analyses were performed cross-sectionally, as repeated measures of lung function were not available in the UK Biobank, and there was an inadequate number of lithium users with longitudinal data in the COPDGene study to perform a meaningful analysis. Statistical significance was defined as two-tailed *p*<0.05.

## RESULTS

In the UK Biobank, we identified 814 individuals using oral lithium and 1,705 individuals using other neuropsychiatric medications (see Methods for full medication list). Compared to users of other neuropsychiatric medications, lithium users were older, more likely to be female and White, had lower BMI, were more likely to be ever smokers but with fewer pack years of smoking, and were more likely to have a diagnosis of mania or bipolar disorder rather than schizophrenia or schizoaffective disorder (**Table 1**).

**Table 1.** A comparison of baseline characteristics between lithium users and users of other neuropsychiatric medications in the UK Biobank.

|  | Lithium users | Other medication users | *p* |
| --- | --- | --- | --- |
| *n* | 814 | 1,705 |  |
| Age, years, mean (SD) | 57.5 (7.8) | 55.3 (8.0) | <0.001 |
| Gender, % |  |  | 0.002 |
| Female | 56.0% | 49.3% |  |
| Male | 44.0% | 50.7% |  |
| Race, % |  |  | <0.001 |
| White | 95.8% | 89.4% |  |
| Other | 4.2% | 10.6% |  |
| BMI, kg/m<sup>2</sup>, mean (SD) | 28.7 (5.6) | 29.5 (5.9) | 0.001 |
| Smoking status, % |  |  | <0.001 |
| Never smoker | 46.6 | 50.6 |  |
| Former smoker | 27.9 | 31.5 |  |
| Current smoker | 25.5 | 17.9 |  |
| Pack years, median (IQR)* | 25.5 (10.8 - 41.2) | 30.0 (17.6 - 46.0) | <0.001 |
| Psychiatric diagnosis |  |  |  |
| Depression, % | 5.8% | 5.1% | 0.48 |
| Mania, % | 4.7% | 2.6% | 0.006 |
| Bipolar disorder, % | 18.1% | 8.9% | <0.001 |
| Schizoaffective disorder, % | 3.1% | 5.5% | 0.008 |
| Schizophrenia, % | 1.8% | 12.6% | <0.001 |

\* In ever smokers

Acceptable or higher quality spirometry data were available for 390 of the 814 lithium users and 757 of the 1,705 users of other neuropsychiatric medications. Within this spirometry subset, lithium users showed similar differences in baseline characteristics vs. users of other medications as in the full cohort, except that the previously observed difference in gender was no longer evident (*p*>0.05). In unadjusted analyses, lithium users had a mean FEV₁ that was 2.6% (± 1.03%) predicted higher than that of users of other medications (**Figure 1A**). This difference remained statistically significant after adjustment for age, sex, race, BMI, and neuropsychiatric diagnoses, including mania, bipolar disorder, schizophrenia, and schizoaffective disorder (**Figure 1B**).

**Figure 1:**
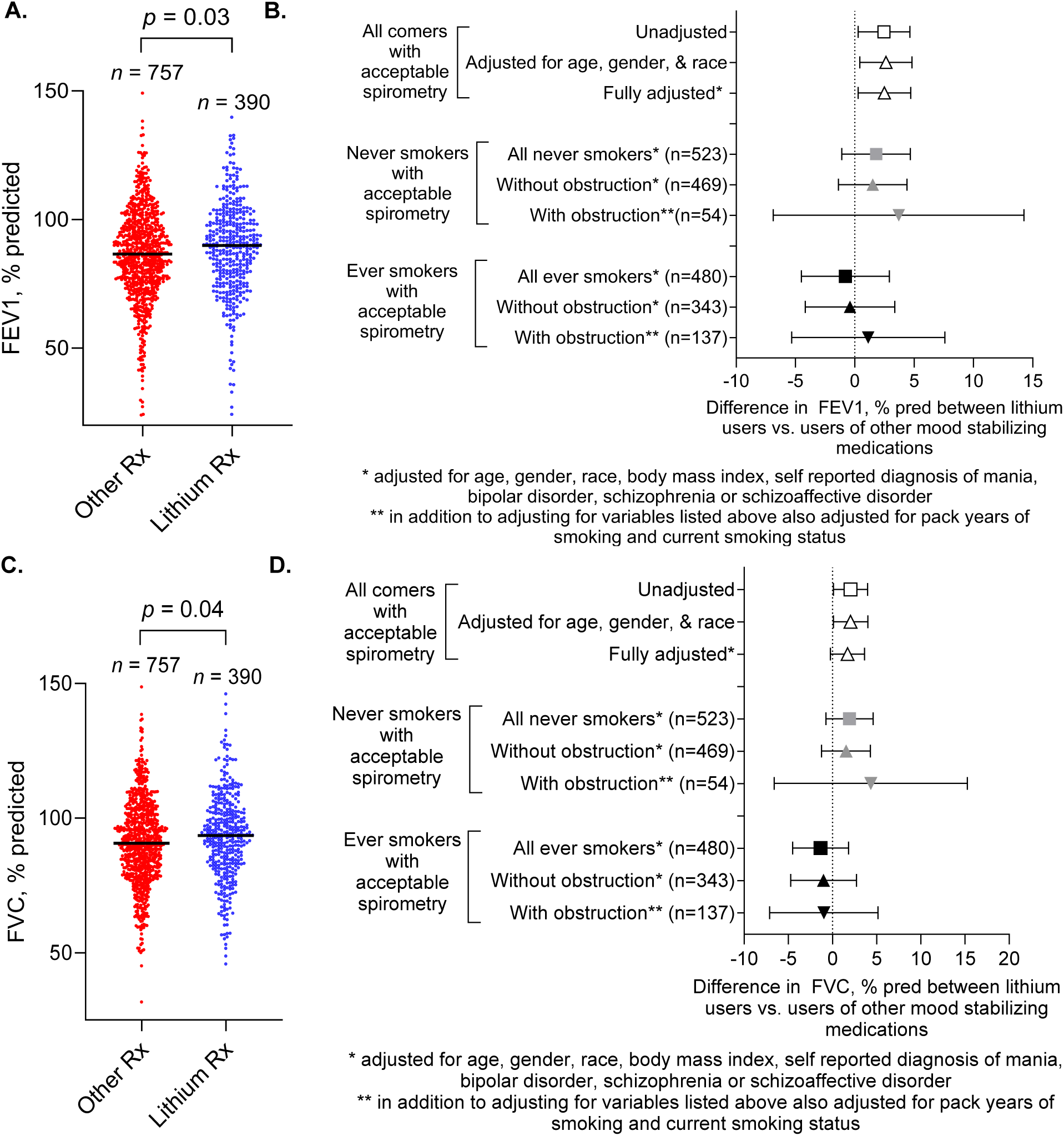
Differences in FEV₁ (% pred) and FVC (% pred) between lithium users and users of other neuropsychiatric medications in the UK Biobank in unadjusted and adjusted analyses across strata of smoking status and airflow obstruction (FEV_1_/FVC<0.70).

To assess the influence of tobacco exposure, analyses were stratified by smoking status (ever vs. never smokers) and further adjusted for pack-years among ever smokers, with additional stratification by the presence or absence of airflow obstruction (FEV₁/FVC < 0.70). In these subgroups, associations between lithium use and FEV₁ were no longer statistically significant, although sample sizes were smaller. Similar patterns were observed for FVC, % predicted (**Figure 1C-D**). No association was observed between lithium use and FEV₁/FVC in either univariate or multivariable models.

Because chest CT-based measures of emphysema were not available in the UK Biobank, we used self-reported MD diagnosis of emphysema as a proxy outcome. Lithium use was associated with a ∼ 50% reduction in the odds of emphysema diagnosis in fully adjusted models, both in all comers and in ever smokers (**Figure 2**). No never-smokers in the cohort had a diagnosis of emphysema, precluding analysis in that subgroup.

**Figure 2.**
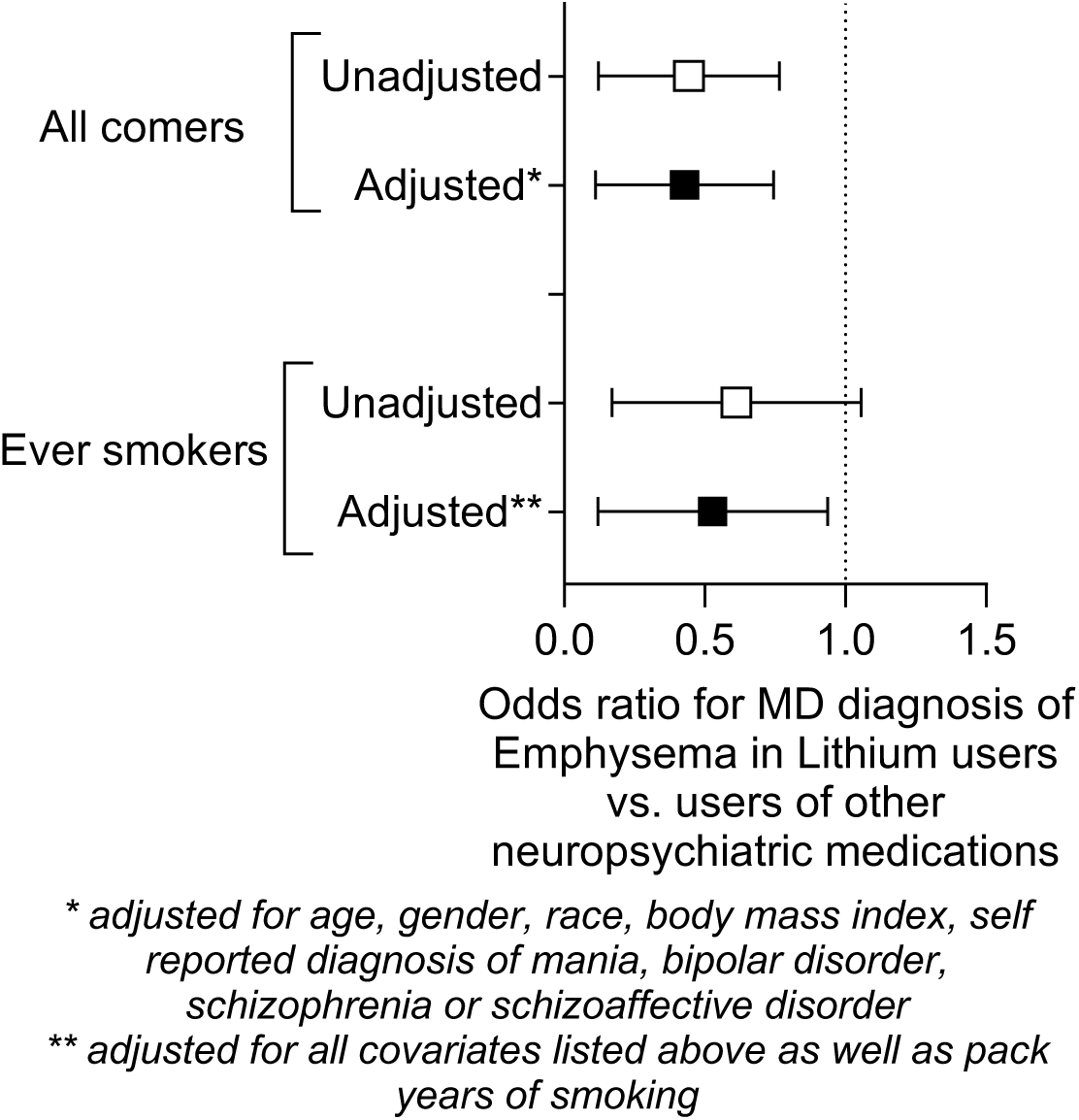
Unadjusted and adjusted odds of self-reported, physician-diagnosed emphysema among individuals using lithium vs. other neuropsychiatric medications in the UK Biobank.

To validate these findings in an independent cohort and to assess the relationship between lithium use and radiographic measures of emphysema, we analyzed data from the COPDGene study. Specifically, we performed a cross-sectional analysis of 45 COPDGene participants using lithium and 1,603 participants using other neuropsychiatric medications. The comparison group included (but was not limited to) users of mood stabilizers also assessed in the UK Biobank, such as olanzapine, quetiapine, risperidone, aripiprazole, and divalproex.

In the COPDGene cohort, lithium users were, on average, younger, had higher educational attainment, and were more likely to be current smokers compared to users of other neuropsychiatric medications (**Table 2**). Lung function measures were higher among lithium users: mean FEV₁ was 8.6 ± 4.0% predicted higher, FVC was 5.4 ± 2.8% predicted higher, and the FEV₁/FVC ratio was 0.05 ± 0.03 points higher compared to users of other neuropsychiatric medications (**Figure 3**). Differences in FEV₁ (% predicted) and FEV₁/FVC remained statistically significant after adjustment for relevant covariates (**Figure 3**).

**Figure 3:**
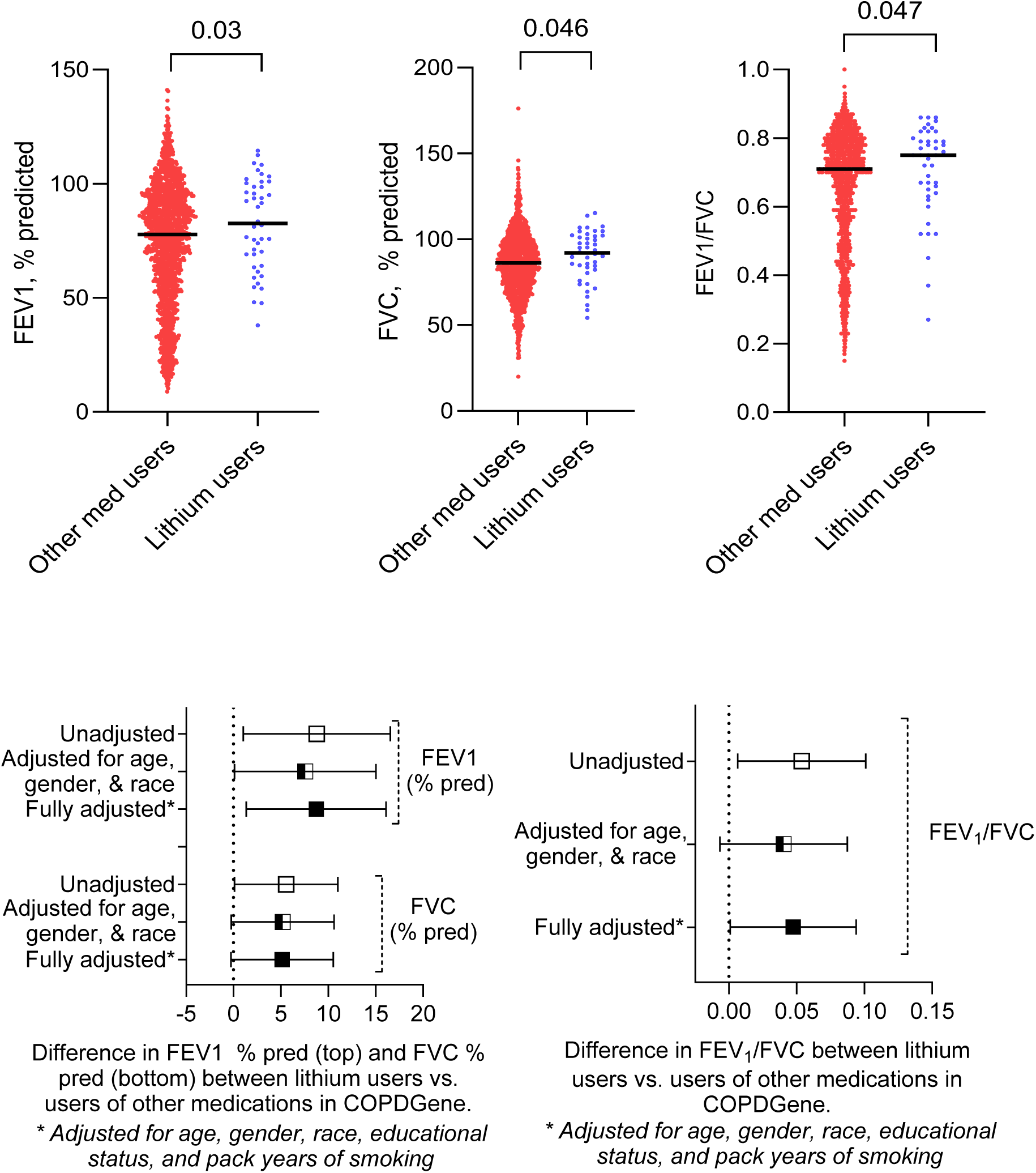
Differences in FEV₁ (% predicted) and FVC (% predicted) between 45 lithium users and 1603 users of other neuropsychiatric medications in the COPDGene cohort in unadjusted and adjusted analyses.

**Table 2:** Baseline characteristics of lithium users vs. users of other neuropsychiatric medications in the COPDGene cohort.

|  | **Lithium users** | **Other medication users** | ***p*** |
| --- | --- | --- | --- |
| *n* | 45 | 1,603 |  |
| Age, years, mean (SD) | 57.2 (8.8) | 59.9 (8.6) | 0.03 |
| Gender, % |  |  | 0.86 |
| Female | 42.2% | 40.9% |  |
| Male | 57.8% | 59.1% |  |
| Race, % |  |  | 0.22 |
| White | 86.7% | 79.1% |  |
| Other | 13.3% | 20.9% |  |
| Educational Status, % |  |  | 0.02 |
| 8<sup>th</sup> grade or less | 8.9% | 2.4% |  |
| High school, no diploma | 13.3% | 9.9% |  |
| High school graduate | 28.9% | 25.2% |  |
| Some college or tech school | 13.3% | 30.4% |  |
| College or technical degree | 28.9% | 22.7% |  |
| Master's or Doctoral degree | 6.7% | 9.4% |  |
| BMI, kg/m<sup>2</sup>, mean (SD) | 30.2 (7.1) | 29.2 (6.7) | 0.30 |
| Pack years, mean (SD) | 44.8 (24.0) | 46.9 (25.4) | 0.60 |
| Smoking status, % |  |  | 0.02 |
| Former smokers | 31.1% | 48.9% |  |
| Current smokers | 68.9% | 51.1% |  |

LAA %F950 was lower, and the average lung density was higher in lithium users vs. non-users (**Figure 4**). In linear regression analyses with % LAA F950 (log transformed due to right skewed distribution) and average lung density as the dependent (outcome) variables, lithium use (yes/no) as the independent (predictor) variable, and age, gender, race/ethnicity, education status, and pack-years of smoking as covariates, lithium use was associated with 0.60 log units lower % LAA −950 (95% CI 0.1-1.1, *p*=0.02), and 8.7 g/L higher adjusted lung density (95% CI 2.3-15.1, *p*=0.008) independent of covariates (**Figure 4**).

**Figure 4:**
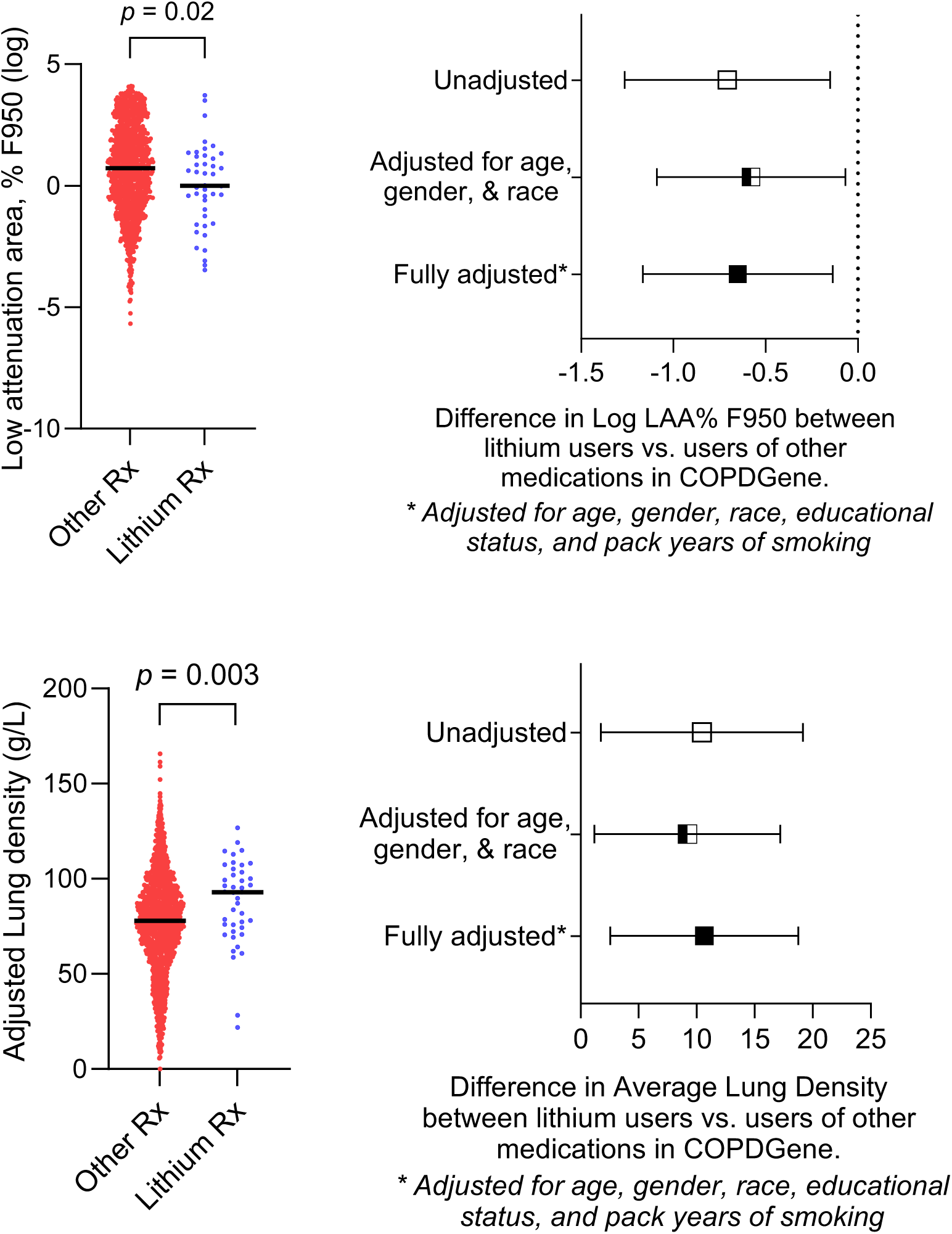
Differences in radiographic low attenuation area (LAA %F950) and average lung density between 45 lithium users and 1603 users of other neuropsychiatric medications in the COPDGene cohort in unadjusted and adjusted analyses.

Next, we performed propensity score matching, pairing each lithium user with two users of other neuropsychiatric medications, adjusting for age, gender, race, education, and smoking history. The average treatment effect (ATE) was estimated with robust standard errors. Balance plots suggested good matching based on similarity in propensity score distributions among matched individuals (**Figure 5**). Lithium use was associated with a 0.63 log-unit lower LAA% F950 (95% CI: 0.29-0.98; *p* = 0.001), and 8.0 g/L increase in adjusted lung density (95% CI: 3.8-12.2; *p* < 0.001, **Figure 6**). We compared results against other causal inference approaches, including nearest neighbor Mahalanobis matching, inverse probability of treatment weighting (IPW), regression adjustment (RA), and doubly robust IPW-regression adjustment (IPWRA), all adjusting for the same covariates using robust variance estimation (**Figure 6**). All methods yielded consistent effect estimates (**Figure 6**), supporting the robustness of our findings.

**Figure 5:**
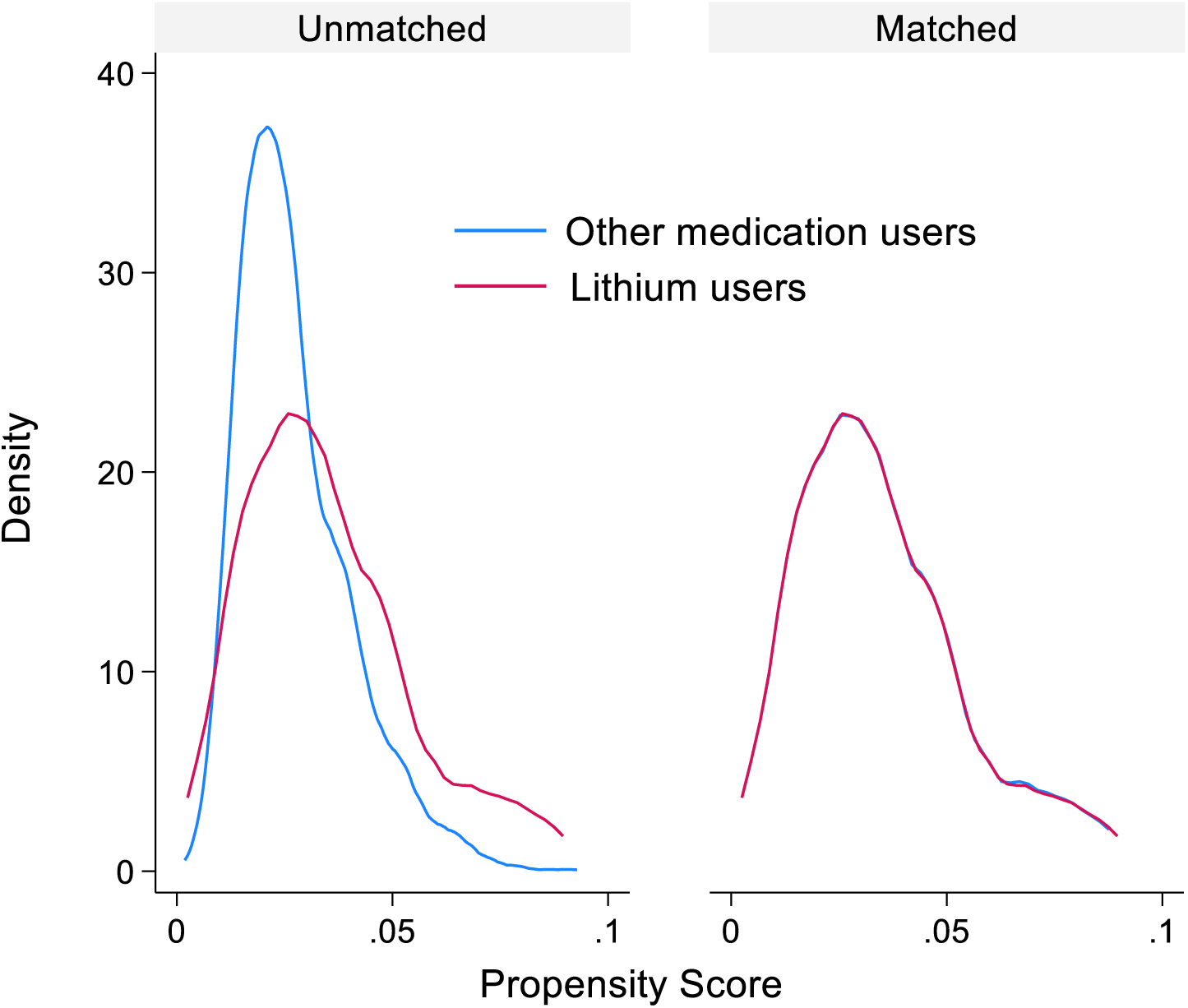
Balance plot depicting histograms of propensity scores in unmatched and matched individuals in the COPDGene cohort.

**Figure 6:**
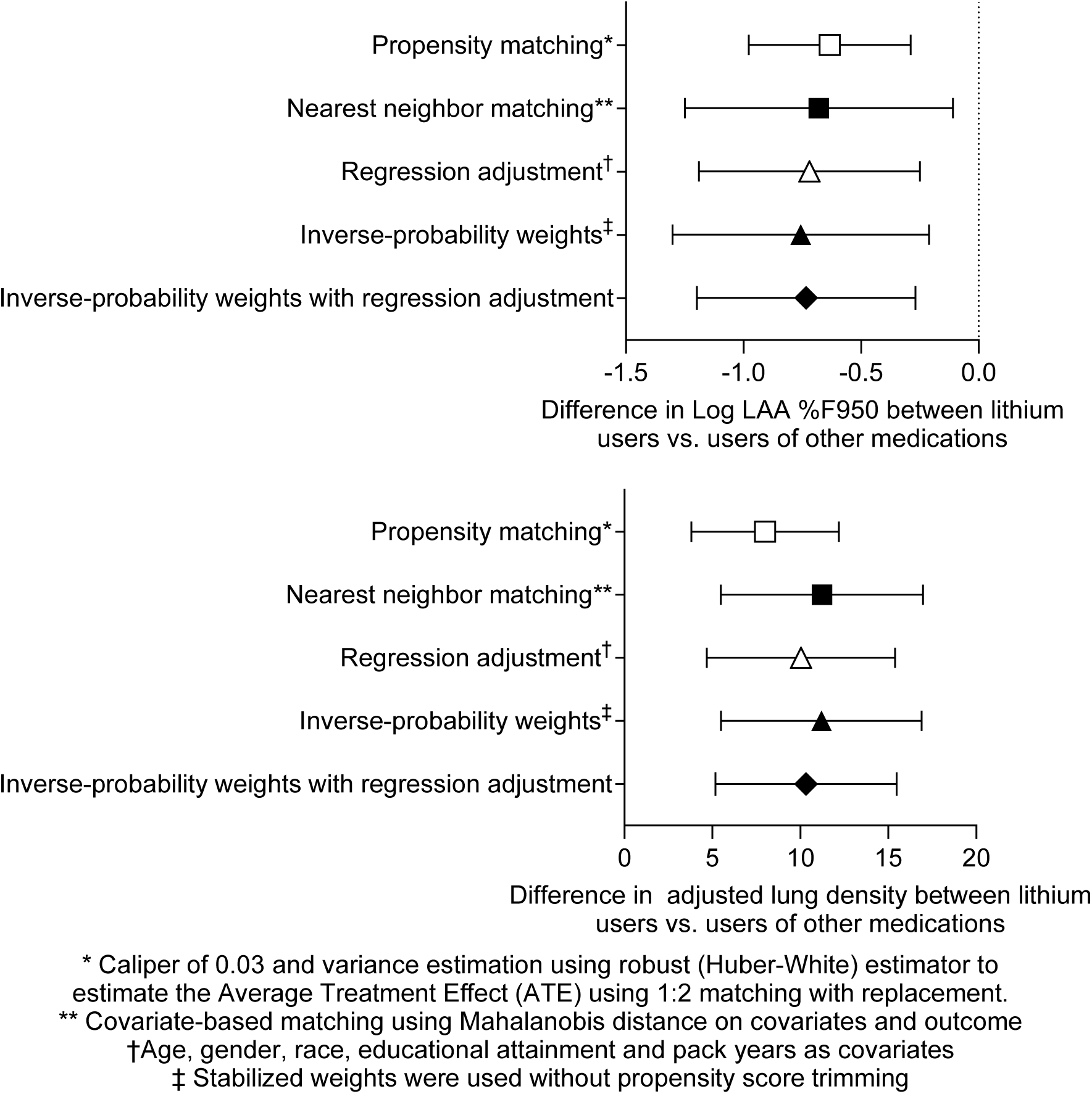
Difference in LAA %F950 (log) and adjusted lung density between users of lithium medication and users of other medications in various types of models in the COPDGene cohort.

Current smoking was not included as a covariate in our models due to the well-described “healthy smoker effect” (21). In COPDGene, current smokers exhibited less emphysema (%LAA F950 1.1 vs. 4.2, *p* < 0.001), higher lung density (86.1 vs. 66.2 g/L, *p* < 0.001), and lower MMRC dyspnea scores (1.3 vs. 1.4, *p* < 0.001) compared to former smokers. This paradox likely reflects the influence of respiratory symptoms on smoking cessation: individuals with more severe symptoms are more likely to quit, making current smoking a potential consequence rather than a cause of milder emphysema. Nevertheless, when we further adjusted our models for current smoking, the association between lithium use and lung density remained statistically significant, while the association with %LAA-950 was slightly attenuated (*p* = 0.07). Including current smoking in the propensity model did not alter the results.

## DISCUSSION

Our findings indicate that the use of the Wnt/β-catenin activator lithium is associated with less emphysema in two independent cohorts. In the UK Biobank, lithium use was associated with higher FEV_1_ and FVC % predicted and a lower likelihood of physician-diagnosed emphysema. In the COPDGene study, lithium users similarly exhibited higher lung function, increased lung density, and reduced radiographic emphysema compared with individuals using other neuropsychiatric medications. These results provide the first human evidence supporting the translatability of our preclinical work, which demonstrated that Wnt/β-catenin activation can mitigate emphysema development *in vivo* and in lung tissue from COPD patients *ex vivo* (1, 2, 22).

Alveolar destruction is a hallmark of emphysema. However, Wnt/β-catenin signaling, which induces alveolar repair, is impaired in emphysema (1–6). This pathway maintains the viability of AXIN2^+^ type 2 cells, which are an evolutionarily conserved alveolar epithelial stem cell (23, 24). During alveolar repair, these cells proliferate in response to Wnt signaling rather than differentiate into type 1 cells (23, 25, 26). In agreement, Wnt/β-catenin enhances the proliferation of type 2 cell organoids (27).

Also, Lithium-induced Wnt activation may have anti-ageing effects. Chronic lithium treatment has been reported to modulate aging biomarkers, such as telomere length and telomerase activity (28). Moreover, the use of lithium medication has been associated with improved longevity in UK Biobank participants (15) and in model organisms such as yeast, *C. elegans*, and *Drosophila* (29). In Japan, higher natural lithium levels in drinking water have been linked to lower all-cause mortality (30). However, other studies suggest that lithium treatment improves healthspan but not lifespan (30). In COPD, we and others have shown that decreased Wnt/β-catenin signaling is associated with early senescence and lung aging (31). The inhibitory effects of lithium on aging-related mechanisms underscore its potential therapeutic benefit for COPD (32).

To our knowledge, this is the first study to report an association between lithium use and less emphysema. Lithium’s therapeutic potential for emphysema merits careful consideration. As an FDA-approved drug with a well-established safety profile, known pharmacokinetics, and low cost, lithium is readily accessible for clinical repurposing. Additionally, lithium may address common COPD comorbidities. Prior studies have suggested that lithium reduces the risk of coronary artery disease (30, 31) and neurodegenerative disorders (32, 33), which are also diseases associated with abnormal Wnt signalling.

However, lithium’s narrow therapeutic window at doses required for mood stabilization (1–2 grams/day) necessitates close monitoring of serum levels. Therapeutic levels range from 5,500 to 8,300 µg/L, typically achieved with daily oral doses of 1–2 g (33). At these levels, multiple adverse effects can occur, including endocrine abnormalities (hypothyroidism and hyperparathyroidism), renal toxicity (chronic kidney disease and nephrogenic diabetes insipidus), neurological effects (tremor and cognitive impairment), gastrointestinal symptoms (nausea, diarrhea, and weight gain), and cardiac toxicity (arrhythmias and bradycardia) (34). Thus, using lithium at mood-stabilizing doses to treat emphysema would require rigorous monitoring of circulating levels with significant risk for adverse effects.

Importantly, our initial *in vitro* data suggest that lithium may activate Wnt signaling at substantially lower concentrations than those required for psychiatric indications (unpublished observation). This raises the possibility that low-dose lithium could confer therapeutic benefit with reduced toxicity. In parallel, several other Wnt/β-catenin activators are under development and may offer improved safety profiles (35, 36). Our current data provide the first human evidence to justify further development and clinical testing of these novel compounds for emphysema.

There are several limitations to our study. The sample size of lithium users in the COPDGene cohort was limited. Therefore, we were unable to stratify our analysis by the presence of airflow obstruction or GOLD stage or to examine the longitudinal progression of emphysema. Other datasets with radiographic emphysema measures, such as SPIROMICS and the Pittsburgh COPD SCCOR, did not include enough lithium users for meaningful analysis. Also, lithium exposure was self-reported rather than confirmed by serum levels. Finally, the duration of lithium use was unknown, which may have influenced the observed associations.

In summary, our study suggests that using the Wnt/β-catenin activator lithium may be associated with less emphysema. These findings agree with our prior *in vivo* and *ex vivo* studies, which have demonstrated that lithium can attenuate emphysema development, further supporting Wnt/β-catenin as a promising novel therapeutic target in COPD. Future research should evaluate the efficacy and safety of lithium for emphysema at lower doses than those used to treat mood disorders. The continued clinical development of other Wnt/β-catenin activators as novel therapies for emphysema is warranted.

## Data Availability

All data produced in the present study are available upon reasonable request to the authors

## ACKNOWLEDGEMENTS

This work was supported by NHLBI grants U01 HL089897 and U01 HL089856 and by NIH contract 75N92023D00011. The COPDGene study (NCT00608764) has also been supported by the COPD Foundation through contributions made to an Industry Advisory Committee that has included AstraZeneca, Bayer Pharmaceuticals, BoehringerIngelheim, Genentech, GlaxoSmithKline, Novartis, Pfizer, and Sunovion. Further, this study is suppoted by R01HL153400 (to DC).

## Notes

### Competing Interest Statement

The authors have declared no competing interest.

### Author Declarations

The IRB at the University of Pittsburgh and at all the study centers where patients were enrolled for this study have approved the research project that resulted in this manuscript.

